# Unveiling Sleep Dysregulation in Chronic Fatigue Syndrome with and without Fibromyalgia Through Bayesian Networks

**DOI:** 10.1101/2025.02.06.25321788

**Authors:** Michal Bechny, Marco Scutari, Julia van der Meer, Francesca Faraci, Benjamin H. Natelson, Akifumi Kishi

## Abstract

Chronic Fatigue Syndrome (CFS) and Fibromyalgia (FM) often co-occur as medically unexplained conditions linked to disrupted physiological regulation, including altered sleep. Building on the work of Kishi et al. [7], who identified differences in sleep-stage transitions in CFS and CFS+FM females, we exploited the same strictly controlled clinical cohort using a Bayesian Network (BN) to quantify detailed patterns of sleep and its dynamics. Our BN confirmed that sleep transitions are best described as a second-order process [14], achieving a next-stage predictive accuracy of 70.6%, validated on two independent data sets with domain shifts (60.1–69.8% accuracy). Notably, we demonstrated that sleep dynamics can reveal the actual diagnoses. Our BN successfully differentiated healthy, CFS, and CFS+FM individuals, achieving an AUROC of 75.4%. Using interventions, we quantified sleep alterations attributable specifically to CFS and CFS+FM, identifying changes in stage prevalence, durations, and first- and second-order transitions. These findings reveal novel markers for CFS and CFS+FM in early-to-mid-adulthood females, offering insights into their physiological mechanisms and supporting their clinical differentiation.

## 1 Introduction

Chronic Fatigue Syndrome (CFS) and Fibromyalgia (FM) co-occur in up to 70% of cases [2]. These conditions share symptoms such as disrupted sleep and exhaustion but have distinct clinical profiles: CFS is characterized by severe, unexplained fatigue worsened by exertion [3], whereas FM is defined by widespread musculoskeletal pain and sensory hypersensitivity [5]. Both conditions disproportionately affect females, with prevalence up to four times higher than in males [4], and are most commonly reported in young to middle-aged adults [3, 5, 9]. They are frequently accompanied by other clinical conditions, including psychiatric and specific sleep disorders [13, 8], complicating the quantification of their underlying effects. Consequently, clinical reviews of existing - mostly observational - studies often lack evidence of their systematic impacts on sleep architecture [8]. The study cohort by Kishi et al. [7] minimized confounding factors and collected polysomnographic (PSG) data from a strictly controlled set of healthy (H), CFS, and CFS+FM females aged 25–55. Exploratory data analysis revealed changes in sleep stage durations and proportions and identified first-order transitions as potential markers enabling clinical interpretation of physiological dysregulation in CFS and CFS+FM.

Recent research in individuals with or without sleep disorders showed that sleep-stage transitions are optimally modelled and analyzed as a second-order process [11, 14]. Leveraging these insights and the CFS/FM dataset [7], we (i) *implement a Bayesian Network (BN) capable of both next-stage prediction and diagnostics*, (ii) *validate the second-order optimality even in a clinical cohort*, and based on that (iii) *identify novel markers for CFS and CFS+FM based on twostage transitions*, providing novel insights into their physiology and supporting their clinical differentiation.

## 2 Materials and Methods

### 2.1 Data

#### Primary Cohort

The data from [7] comprises PSG recordings from 52 females, carefully selected to ensure homogeneity and avoid confounding. The cohort included 26 healthy controls (H, aged 38 *±* 8 years), 14 individuals with CFS only (aged 37 *±* 9 years), and 12 individuals with CFS and FM (CFS+FM) (age: 41 *±* 6 years). Rigorous exclusion criteria were applied, including the presence of clinically evident sleep disorders or other psychiatric conditions. Subjects also refrained from alcohol, caffeine and strenuous activities before the study, and menstruating individuals were evaluated during the follicular phase of their cycles. The PSG data were recorded during a single night in a controlled hospital environment, with sleep stages scored every 30 seconds. This carefully curated data set enables robust estimation of the underlying effects of CFS and CFS+FM in early to mid-adulthood females.

#### Validation Cohorts

The *Bern Sleep–Wake Registry* (BSWR) from the University Hospital Bern and the open-access *Sleep Heart Health Study* (SHHS) are clinical and general-population data sets used to assess the robustness and validate the next-stage predictions of our developed model. To ensure demographic alignment with the primary cohort, subsets of 834 and 1227 females aged 20–60 were selected from the BSWR and baseline-SHHS (SHHS1), respectively. The BSWR challenged the model’s predictive capabilities with a population of sleep-disordered subjects, while SHHS1 assessed it in a general population.

To ensure consistency across analyses, sleep-scoring in all data sets was standardized to five sleep-wake stages following the AASM guidelines [1]: W = Wake, R = Rapid-eye-movement sleep, and (N1, N2, N3) non-R sleep-states.

#### Preprocessing

Having a controlled homogeneous study population (females of the same age), we considered Health Status (HS): H, CFS, CFS+FM, as the only demographic variable. When modelling sleep dynamics, we ignored the PSG recordings before the first non-W stage. We identified continuous bouts (runs) of each stage—denoted *S*_*t*_, indexed by *t*—and recorded their durations (*D*_*t*_). This reduced the original 44,581 sleep-stage-epochs to 7,254 bouts. For each bout, we also recorded the time-since-sleep-onset (*T*_*t*_, TSSO) and cumulative characteristics (*C*_*t*_) monitoring either sleep-time (CST=N1+N2+N3+R) or restorative-sleep-time (CRST=N3+R). To utilize the existing Bayesian inference implementation [12], we discretized the TSSO into five 90 minutes categories (*<*90, 90-180,…, *>*360) of expected sleep cycles and split *C*_*t*_ and *D*_*t*_ variables into four groups based on (25, 50, 75)%-quantiles, with the possible additional class of 0, if present in the corresponding variable. The validation cohorts underwent the same preprocessing, yielding 113,071 and 150,296 bouts, respectively.

### 2.2 Bayesian Networks to Capture Sleep Stage Dynamics

A Bayesian Network (BN) is a statistical framework that encodes probabilistic relationships between variables and can represent cause-effect relationships under additional causal assumptions [12]. These relationships can be learned from data (structure learning), defined by experts (incorporating domain knowledge), or by combining both approaches. Represented as a Directed Acyclic Graph (DAG), BNs offer several advantages, including reduced parameter complexity and interpretable predictions—a critical requirement in the healthcare field.

A compelling feature of BNs is their ability to fix specific nodes (variables) at desired levels, such as the health status (HS) to H or CFS, representing what is referred to as an *intervention*. This enables do-calculus and the simulation of causal counterfactuals [10], addressing what-if questions such as ours of *how sleep patterns change if a healthy individual were to develop CFS or CFS+FM*. Dynamic BNs (DBNs) extend the approach to temporal processes by incorporating dependencies across time, including lagged features. This makes them particularly suited for modelling sleep transitions.

#### Experimental Setup

Rather than relying only on data-driven structure learning algorithms, we dependencies using expert knowledge, as shown in Fig. 1. This included mandatory (solid) edges encoding the impact of all previous stages on the following ones (in red) and the impact of HS on *S, C, D* (in green). Similarly to HS, the possible (dashed) impact of TSSO (*T*) on (*S, C, D*) in yellow was considered. Further, we hypothesized that transitions in *S* might be better explained by considering cumulative sleep variables *C*, which naturally depend on *T*. Existing work suggested that including stage-duration *D* (depending on *S* as red-dashed edges) might boost next-stage predictions (blue-dashed edges) [14]. To systematically evaluate the impact of each variable’s inclusion (including BN-lag/order), we used linear regression associating BN-performance (to identify next-stage and HS) with indicators of each variable’s inclusion and the lag used.

**Fig 1.**
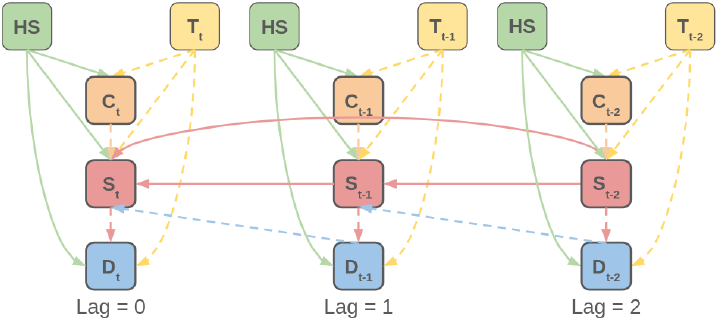
Illustration of the full-structure Bayesian Network of lag = 2. HS = health status (healthy, CFS, or CFS + FM), *T_t_* = time since the sleep onset, *C_t_* = cumulative sleep, *S_t_* = sleep-stage, *D_t_* = duration of sleep-stage, chronologically indexed by *t*.

Clinically, beyond quantifying the effects of CFS and CFS+FM (HS-node), this allowed us to test whether the cumulative sleep (CST, CRST) better explains sleep dynamics than the TSSO. Despite the well-known effect of TSSO on sleep macro-architecture (e.g., higher R-% in the second half of the night), its influence on sleep dynamics remains inconclusive [14].

## 3 Results

### 3.1 Descriptive Statistics

**Traditional sleep variables** of the primary dataset are described in detail in the original work, which also reports their Tukey-Kramer multiple comparisons concerning the HS [7]. The significant differences were identified for the total sleep time [mins] (H > CFS), N1 and N2 [mins] (H > CFS+FM), N3 [mins] (CFS+FM > H), and REM [mins] (H > CFS), c.f., Table 2 in [7].

**Occurence of stage-specific bouts and their duration** is presented in Table 1. H experienced significantly more R-bouts than both CFS conditions and more N1-bouts than CFS+FM. In addition, CFS+FM exhibit more N3-bouts than CFS only. The subject-aggregated means of stage-specific bout durations did not exhibit significant differences across HS.

**Table 1.** Mean (SD) bout statistics for H, CFS, and CFS+FM subjects. Bouts indicate the average number of stage runs experienced, and Duration their length in minutes. Significant pairwise comparisons according to the Tukey-Kramer procedure are marked with * and ** for p-value < (0.05 and 0.01), respectively.

| Stage | Characteristic | H | CFS | CFS+FM | Significant Pairs |
| --- | --- | --- | --- | --- | --- |
| W | Bouts | 22.5 (6.4) | 22.5 (8.5) | 19.7 (5.6) | - |
|  | Duration | 2.4 (1.5) | 3.6 (3) | 3.2 (2.1) | - |
| N1 | Bouts | 44.6 (17.2) | 37.5 (17.7) | 29.8 (8.9) | H - (CFS+FM)* |
|  | Duration | 1 (0.2) | 1 (0.2) | 0.9 (0.2) | - |
| N2 | Bouts | 50.2 (16) | 43.1 (11.9) | 52.7 (14.8) | - |
|  | Duration | 5 (2) | 5.1 (1.7) | 4.1 (2.1) | - |
| N3 | Bouts | 18.4 (10.2) | 15.6 (6.6) | 25.1 (12) | CFS - (CFS+FM)* |
|  | Duration | 2.2 (2.2) | 2.8 (1.9) | 3.3 (2) | - |
| R | Bouts | 13.4 (7) | 6.5 (4.8) | 8.2 (3.8) | H - CFS**; H - (CFS+FM)* |
|  | Duration | 8.2 (5.8) | 12.6 (8.5) | 10.8 (7.9) | - |

**Table 2.** The impact of BN-included variables on the performance metrics. Significant variable associations and model explanations based on F-test are highlighted as p-value < 0.05, 0.01, and 0.001, respectively. The 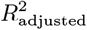 (not tested) and F-statistic refer to regression models evaluating the systematic impact of included variables on the performance metric across different BN-settings and not to any specific BN.

| Variable | Accuracy $[S_t]$ | F1-score $[S_t]$ | AUROC $[HS]$ |
| --- | --- | --- | --- |
| lag = 0 | 44.00 | 50.84 | 71.59 |
| lag = 1 | 68.03 | 72.75 | 74.13 |
| lag = 2 | 72.08 | 73.07 | 74.29 |
| lag = 3 | 68.91 | 70.05 | 75.63 |
| lag = 4 | 64.78 | 65.94 | 75.85 |
| TSSO | -4.14 | -4.62 | -4.14 |
| Stage-Duration | -3.29 | -3.87 | 2.45 |
| CST | -1.53 | -1.65 | -3.98 |
| CRST | -12.03 | -12.56 | -8.42 |
| Model's F(9, 51) | 1252.09 | 2323.33 | 4715.47 |
| Model's $R^2_{\text{adjusted}}$ | 0.995 | 0.997 | 0.999 |

### 3.3 Structure Identification

The selection of the optimal structure was guided by a computational experiment in which each variable (except mandatory *S*_*t*_ and HS) along with its associated dependencies (edges), as depicted in Fig. 1, was either included or excluded from the DAG. This evaluation used HS-balanced 3-fold cross-validation (CV) with subject-wise splits, allowing performance quantification for each variable combination while ensuring a reasonable number of subjects were included in the testing fold. The performance metrics used included: **next-stage accuracy** and **F1-score**, and **average AUROC** (of AUROCs specific to H, CFS, and CFS+FM). The results are summarized in Table 2. Based on next-stage performance metrics, we identified lag=2 as optimal, confirming [14], as both accuracy and F1-score were the highest under lag=2 and appear to decrease with larger lags. The AUROC, indicating capability to identify HS, was up to 1.56% better for higher lags, but their consideration would lead to an expected decrease of up to 7.3% in accuracy/F1-score. All TSSO, CST, and CRST yielded a systematic decrease in all performance metrics. This may suggest that sleep dynamics and HS identification are either unrelated to these variables or that the BN was under their inclusions over-parametrized, as the number of parameters to predict the *S*_*t*_ just from *S*_*t−*1_, *S*_*t−*2_, HS involves 75 = 5 *×* 5 *×* 3 parameters which scale by 4-5 with inclusion of every additional (TSSO, CST, CRST) variable. Based on that, we chose the BN of lag = 2 with included stage durations as the final model to demonstrate the CFS and CFS+FM effects. Despite slightly reduced next-stage predictive accuracy due to Duration inclusion, this model seems to significantly enhance the identification of HS. Our evaluations tried to find a compromise between the best performance in the next stage and diagnosis identifications.

### 3.3 Performance and Generalization

The final BN (lag = 2, including stage durations) achieved 70.61 (1.9)% and 69.2 (2.7)% in mean (SD) on-subject next-stage accuracy and F1-score, and the HS AUROC of 75.36 (8.3)%. For each subject, the HS probabilities were averaged over all *S*_*t−*2_ *→S*_*t−*1_ *→S*_*t*_ sequences available.

To further test the robustness of the final BN to capture sleep dynamics, we evaluated its predictive accuracy on BSWR and SHHS1. Despite training on a small sample of 52 strictly controlled subjects, BN achieved 69.78 (7.25)% and 60.1 (11.62)% in mean (SD) on-subject accuracy, 70.94 (9.1)% and 59.83 (11.56)% in on-subject F1-score, on BSWR and SHHS1, respectively. Considering that both test data sets represent out-of-domain samples from general and clinical cohorts, respectively, with considerable domain shifts, these results suggest the particular robustness of our BN. In contrast, similar work reported 62.2% testing accuracy (corresponded to in-domain cross-validation assessment) on a broad sample of 3,202 PSG recordings with excluded sleep-disorders [14].

### 3.4 Effects of CFS and CFS+FM via Interventions

We evaluated three interventions by fixing the HS node of our final BN to H, CFS, and CFS+FM levels, allowing sampling from arbitrary nodes under specified conditions. Assuming no hidden confounding, which is reasonable in our strictly controlled cohort, comparing samples for CFS-vs-H and (CFS+FM)-vs-H enables estimating the causal effects of the two conditions. Arbitrary 95% credible intervals (CI) were constructed by generating 1,000 *×* 1,000 samples and calculating median (= estimate) and (2.5, 97.5)%-quantiles (= CI-bounds).

#### Bouts Duration

Fig. 2 presents BN-based CIs for expected stage durations. Discretized *D*_*t*_ levels were represented by mid-points and multiplied by obtained samples. Both CFS and CFS+FM exhibit prolonged W and N3 durations, indicating reduced sleep efficiency and increased physically-restorative drive. CFS additionally exhibits extended R stages, linked to cognitive restoration, despite fewer R bouts. In contrast, CFS+FM shows shorter N1 durations, likely compensating for increased W and N3. Notably, CFS does not display reduced durations in any stage, suggesting compact sleep despite decreased efficiency.

**Fig 2.**
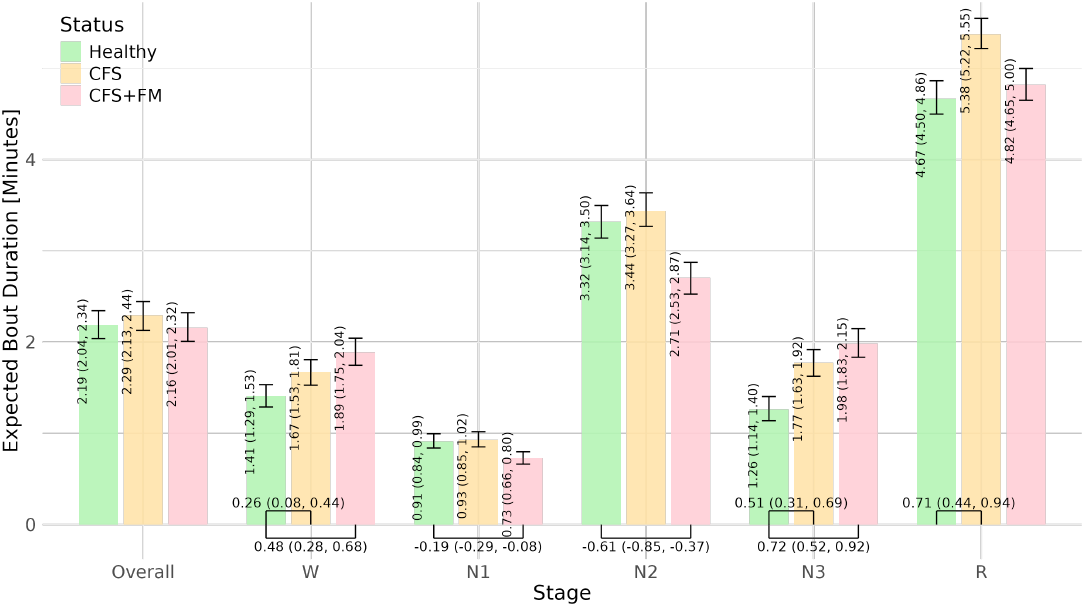
Expected durations of sleep-stage-bouts for H, CFS, and CFS+FM groups, presented with 95% CI as vertical error bars. Horizontal brackets indicate significant differences between conditions, accompanied by their estimates and 95% CI.

#### First-order transitions

*S*_*t−*1_→*S*_*t*_ expected for H are shown in Fig. 3.a and the CFS and CFS+FM effects in Fig. 3.(b-c). The effects were quantified without conditioning on any particular stage and describe the overall sleep dynamics. Below, we write in **bold** alterations by at least 10%. CFS showed reduced R% and increased N1→W, **R→W** that were compensated by decreased **N1**⇄**R**. The changes were more pronounced in CFS+FM, which showed increased (N2, N3)% and decreased (N1, R)%. Further, CFS+FM exhibited significantly increased (W, **N1, R**)→**N2**, N2→N3, N3→(W, N1), and decreased **(W, N2, R)→N1, N1→R**, and **N3→N2**. Our findings confirm all alterations found by [7] in their Fig. 1. We additionally identified increased N1→W (c.f., [6]) in CFS (compensation for decreased N1→R) and disruptions in N2 for CFS+FM [7].

**Fig 3.**
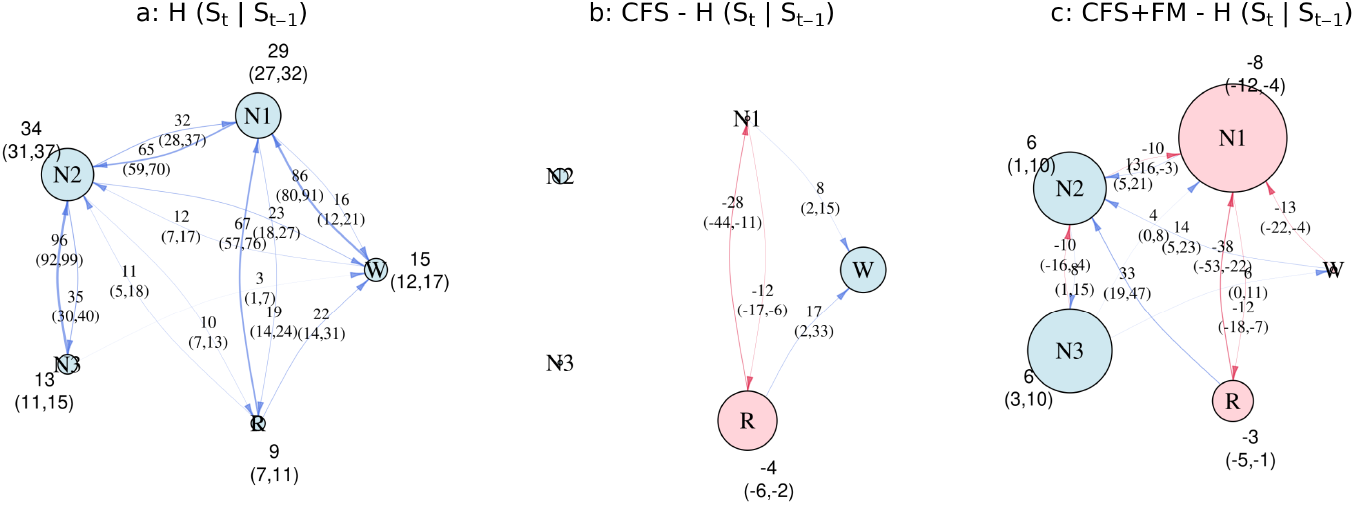
Lag-1 sleep-stage transition dynamics for Healthy (H), Chronic Fatigue Syndrome (CFS), and CFS with Fibromyalgia (CFS+FM). Panel (a) illustrates the expected transitions for H, with node sizes proportional to the prevalence of *S*_*t*−1_ stages and edges indicating transition (*S*_*t*−1_ → *S_t_*) probabilities. Panels (b) and (c) depict the differences in stage prevalence and transition probabilities due to CFS and CFS+FM, in comparison to H, respectively. Positive and negative values are shown in blue and red, respectively, and significant alterations are annotated with their estimates and 95% credible intervals.

#### Second-order transitions

*S*_*t−*2_→*S*_*t−*1_→*S*_*t*_ in Fig. 4 provide deeper insights into sleep dynamics. The first row represents expected transitions for H, while rows 2 and 3 depict the effects of CFS and CFS+FM. Each column (a–e) corresponds to a different starting stage *S*_*t−*2_. In some cases, the alterations are only in *S*_*t−*1_ (nodes), or follow-up transitions (*S*_*t−*1_→*S*_*t*_, edges), both conditioned on *S*_*t−*2_ and extending the unconditioned first-order results from Fig. 3.

**Fig 4.**
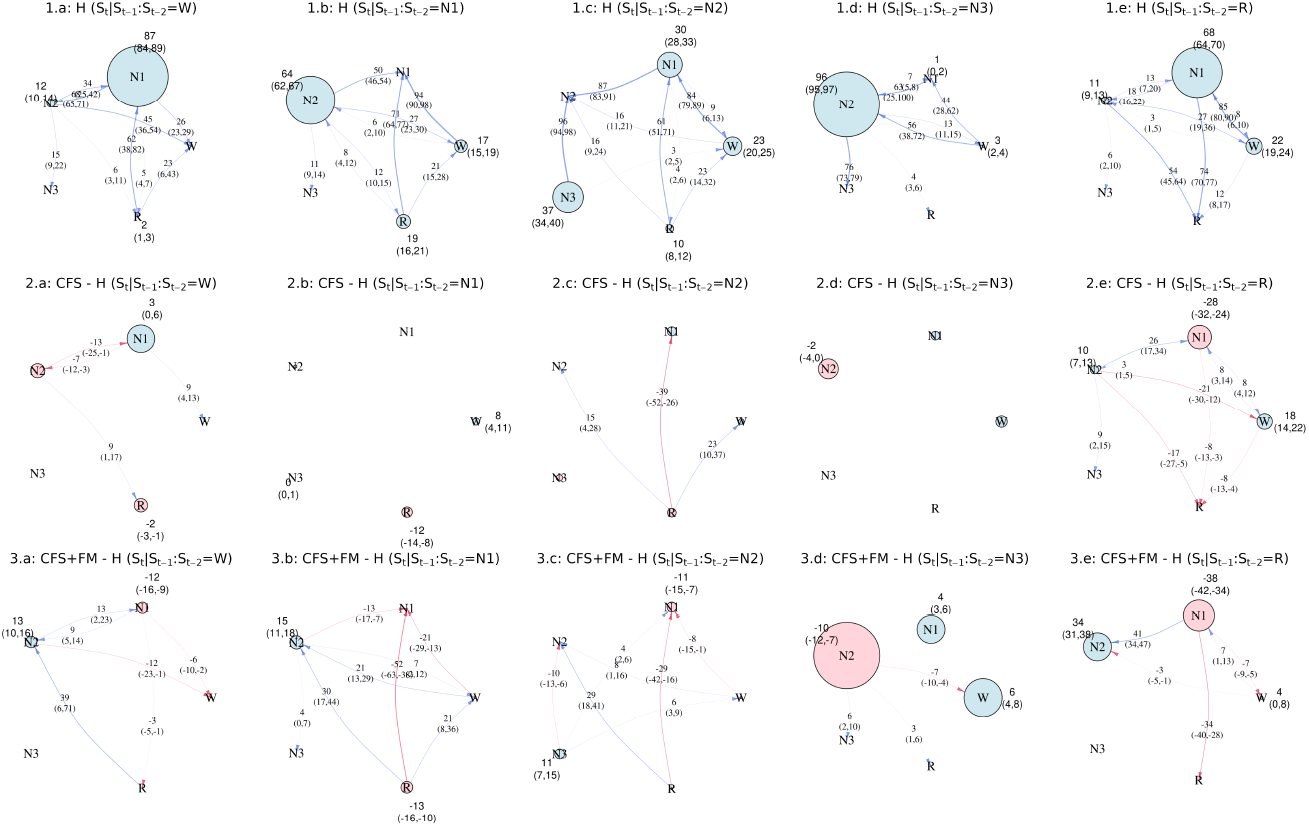
Lag-2 sleep-stage transition dynamics (*S_t_* ← *S*_*t*−1_ ← *S*_*t*−2_) for Healthy (H), Chronic Fatigue Syndrome (CFS), and CFS with Fibromyalgia (CFS+FM). The first row shows the expected dynamics for H, while rows 2 and 3 display their changes due to CFS and CFS+FM, in comparison to H, respectively. Node sizes represent *S*_*t*−1_ prevalence (or its difference), and edges illustrate transition probabilities (*S_t_* ← *S*_*t*−1_) or their differences. Positive and negative values are shown in blue and red, respectively, and significant alterations are annotated with estimates and 95% credible intervals.

In ***CFS***, key disruptions included increased **R→W** (with subsequent increases in N1 and decreases in R) and **R→N2** (followed by increased **N1**, N3, and decreased **W, R**), along with reduced **R→N1**. These patterns suggest an impaired ability to achieve or maintain restorative R sleep, compensated by nonrestorative transitions within light sleep (N1, N2). Additionally, W→R, common in healthy individuals during the second half of the night, was decreased and supplemented by W→N1. More frequent N1→W, at the expense of **N1→R**, further contributed to reduced sleep-efficiency and increased fragmentation.

In ***CFS+FM***, disruptions included increased R→W (with subsequent increases in N1 and decreases in N2) and **R→N2**, along with reduced **R→N1** (followed by decreased W and **R**, and increased **N2**). Particularly increased **N2→N3** (followed by increased transitions to W and N1, and reduced to **N2**), reflecting a compensatory drive for deep sleep (N3) likely linked to FM’s restorative needs, while also indicating difficulty maintaining smooth sleep cycling. Reduced **W→N1** and increased **W→N2** suggest a shift towards intermediate sleep stages at the expense of lighter sleep, possibly as a response to pain-related disruptions. Increased awakenings from N3 (compensated by reduced **N3→N2**) and from R further destabilized transitions between restorative and lighter stages, amplifying sleep fragmentation and reducing efficiency. These findings align with FM’s symptomatology, where widespread pain increases the need for deep sleep (N3) but disrupts restorative sleep transitions, highlighting the need for tailored treatments to improve both sleep and pain management.

## 4 Discussion

In this study, we constructed a Bayesian Network (BN) to quantify the effects of Chronic Fatigue Syndrome (CFS) and its interaction with Fibromyalgia (FM) on sleep dynamics. Using a strictly controlled dataset [7], we confirmed that second-order transitions optimally describe sleep patterns, extending findings from non-clinical populations [14]. Despite a relatively small dataset of 7,254 bouts from 52 subjects, our BN achieved robust next-stage predictions with in-domain (out-of-domain) accuracies of 70.6% (60.1–69.8%), respectively. This capability enabled the successful differentiation of healthy (H), CFS, and CFS+FM groups (AUROC: 75.4%), showcasing sleep dynamics’ potential for diagnostics. Based on that, we used interventions to quantify the effects of CFS and CFS+FM compared to H on different aspects of sleep dynamics.

Both conditions exhibited prolonged wakefulness (W) and N3 stages, reflecting reduced sleep efficiency (aligning with insomnia-symptoms in CFS [13]) and increased physical restoration needs, particularly pronounced in CFS+FM. Additionally, CFS showed extended R durations related to an increased sympathetic activity and a higher need for cognitive restoration, while CFS+FM demonstrated reduced durations of N1 and N2. Interestingly, the duration of any stage did not decrease in CFS, suggesting that their sleep despite reduced efficiency - may remain relatively compact.

First-order transitions confirmed all previous findings [7], and - thanks to the joint estimation of transition-probabilities in our BN (as opposed to the pairwise comparisons in [7]), revealed three additional compensatory transitions. CFS is marked by frequent and prolonged awakenings from the N1 and R stages, disrupting “healthy” oscillations between them. This suggests reduced sleep efficiency at the expense of R sleep, potentially contributing to fatigue from both, insufficient sleep quantity and inadequate autonomic or cognitive restoration. In contrast, CFS+FM is characterized by awakenings from deep N3 sleep, into which they tend to transition more frequently. FM, associated with physical pain and discomfort [5], appears to drive both the increased N3 duration and the pressure to transition to N2 instead of N3 across stages.

The second-order transitions provided a novel and detailed perspective on sleep alterations. Both conditions exhibited increased transitions into W, particularly from R, reflecting reduced sleep efficiency. For CFS, fewer alterations were observed, consistent with their longer bouts. The results highlighted CFS-specific patterns of awakenings from N1 and R, difficulties maintaining R (due to transitions into N2), and challenges achieving R. These disruptions may represent the patients’ common complaint of “unrefreshing sleep”, either as a cause or a consequence of fatigue, as commonly reported in CFS. In contrast, CFS+FM showed more widespread alterations, including frequent awakenings from both R and N3, coupled with a marked compensatory drive to achieve and sustain N3, likely driven by the physical symptoms of FM.

## Conclusions

Our study confirms that sleep transitions are best described as a second-order process, even in diseased clinical subjects. Using a strictly controlled cohort of young-to-middle-aged females, we identified the effects of CFS and CFS+FM on alteration sleep and its dynamics, supporting their clinical differentiation. These findings highlight the potential of sleep dynamics as a noninvasive diagnostic tool and may suggest differing therapeutic needs tailored to the unique sleep disruptions observed in these conditions. Our findings should not be directly generalized to males and older subjects, as our study population did not include them, necessitating further evaluations in these groups.

## Data Availability

All data produced in the present study are available upon reasonable request to the authors

